# Reproducibility Policies in Cardiology Journals: The REPLICA Cross-Sectional Study

**DOI:** 10.1101/2022.08.04.22278423

**Authors:** Lucas Helal, Filipe Ferrari, Danielle B Rice, Nadera Ahmadzai, Becky Skidmore, Daniel Umpierre, David Moher

## Abstract

**Importance:** Transparency and data sharing are valuable practices in research, contributing to improved precision and flexibility in cumulative evidence; and ultimately expanding the research ecosystem by addressing one of the philosophical research norms that implies that knowledge belongs to society.

**Objectives:** The objective of the Reproducibility Policies In Cardiology Journals (REPLICA) study was to estimate the proportions of policies and guidance for reproducibility and transparency practices among Cardiology journals, as well as to determine details of completeness of reporting and data sharing conditions whenever disclosed.

**Design:** Cross-sectional analysis.

**Setting:** Cross-sectional study through analyses of journals deposited in the National Library of Medicine (NLM) Catalog tagged with the “*Cardiology*” and “*Vascular Diseases*” entry terms.

**Eligibility Criteria:** Cardiology journals from the NLM Catalog database that published at least one randomized clinical trial in 2018. Journals that published articles in English, Spanish, French, or Portuguese and were available in MEDLINE/PubMed were eligible for inclusion.

**Exposures:** The exposures were mainly related to journal’s characteristics such as publisher operations characteristics (e.g., journal access only by subscription), indexation in the DOAJPlus, requirement for registration for RCTs, and others.

**Main outcomes and measures:** We prespecified a primary composite outcome composed of data-sharing policy or guidance. Secondary outcomes were proportions of reporting guidelines within the journal’s instructions for the author’s section (e.g., CONSORT), and also other components of sharing practices.

**Results:** We assessed 148 journals. Of them, 74 (50.0%, 95%CI 41.9% to 58.1%) presented policy or guidance for data sharing. We found guidance for data sharing in 68 journals (47.5% 95%CI 39.4% to 55.8%). Notably, among them, only two mentioned sharing individual participant data (IPD). Regarding guidelines for article reporting, we identified that 132 journals displayed guidance for authors, in which 27 (20.45%, 95%CI 14.34% to 28.29%) had CONSORT and EQUATOR Network guidance content.

**Conclusion and relevance:** In summary, we found a mild proportion of policies and guidance for data-sharing. Moreover, transparency practices inclined to RCTs are suboptimal, as mirrored by the very low prevalence of IPD data-sharing policy and guidance as well as specific reporting guidelines instructions for RCTs.

**Key Points:** *Question:* What is the proportion of journals displaying policies and guidance about data sharing in cardiology journals?

*Findings:* We found a low prevalence of policy and guidance for data sharing in Cardiology journals, as well as transparency and reproducibility practices; details, individual participant data sharing, registration, and completeness of reporting, for example.

*Meaning:* Journals play a role in driving reproducibility and transparency among scientific areas. Stakeholders involved in the editorial processes should be open to understand the valuable impact of data-sharing practices and learn how to implement such mechanisms, that being the case.

## Rationale and Background

Data sharing may reduce publication bias, substandard dissemination of studies, and retractions.^1-3^ The importance of data sharing can be considered in the context of harms identified in trials. For example, harms experienced by research participants can be investigated through deidentified data that is shared by study authors.^4^ And, beyond the impact of the risk-benefit ratio in which a research participant is exposed (and previously known by investigators), it is often not sufficient to improve data-sharing practices spontaneously for that púrpose.^5^ Academic journals are the gatekeepers for a sustainable good science, despite some ongoing caveats.

Journals can contribute to replication failure. Many of them lack editorial policies and guidance for replication^6^ whereas the International Committee of Medical Journal Editors (ICMJE) released a statement for their journals in 2019 requiring a mandatory disclosure in the manuscripts whether authors would share or not share their data for randomized clinical trials (RCTs) – and it should be clearly stated in an journal editorial policy.^7^ Independent initiatives like the Center for Open Science (COS) drive data sharing by providing workflows for sharing specific items (e.g., the Transparency and Openness Guidelines/Factor – TOP).^8^ There are also strong data-sharing standards such as the one adopted by the BMJ, which operates with a tiers beyond then “*strongly encouraged*".

Cardiology RCTs seem to have a low prevalence of data sharing,^5^ similar to other disciplines in biomedicine.^9,10^ Inadequate data sharing in journals might reflect a lack of guidance and/or policy mandates by journals and publishers. The lack of transparency about editorial instructions is a problem. We should recognize that instructions for open and transparent science are still incipient. This should be emphasized given the nature of the process as a facilitator of study’s reproducibility and replicability. For example, an experiment attempting to reproduce 100 independent studies conducted by Nosek and collaborators was only able to achieve success on reproducibility of results in 47% of studies, even tough working by the side of the study’s original authors.^11^

Therefore, our primary objective was to estimate the proportion of a composite outcome on data sharing (editorial policy or guidance) in Cardiology for RCTs. Secondary objectives were estimates of prevalences of reporting guidelines instructions, sharing practices details, open-access level, and other variables related to transparency/reproducibility that are accountable to data-sharing practices.

## Methods

This manuscript was written based on the Strengthening the Reporting of Observational Studies in Epidemiology Statement (STROBE Statement) whenever possible.^12^ In a public repository, a full protocol, with study materials, was deposited prior to data collection (*repository link omitted for peer-reviewing purposes*). As an operational concept, for “*has policy*’ we considered any content in the Aims, Scope and Instruction for Authors towards the data-sharing mandate as a whole in the journal. For “*has guidance*", among those which “*has policy*", it is about details on how to share the data and other contents and not only declare that the journal adheres to such practice.

### Eligibility criteria

To be eligible for the study, we considered journals classified as “*Cardiology*” of “*Vascular Disease*” by the National Library of Medicine (NLM) Catalog, and published at least one RCT in 2018. Journals without websites in English, Spanish, Portuguese or French languages were excluded.

### Electronic searches and eligibility process

An experienced medical information specialist (BS) queried the NLM Catalog through Ovid© to search for eligible journals. We used the broad subject terms “*Cardiology*” and “*Vascular Diseases*” in the NLM Catalog, considering the database as fully accurate at the journal level.

We then ran searches for each journal in PubMed/MEDLINE© with the Cochrane Highly Sensitive Search Strategy (HSSS) filter for RCTs through Ovid©,^13^ limiting publications from January 1, 2018, to December 31, 2018. Thereafter, a library in Zotero (Zotero, v. 5.0.76) was created with all retrieved journals that had at least one publication with all 2018 respective publications. One reviewer (FF) screened titles and abstracts of journal’s reports. Whenever one RCT was detected as published in 2018, the journal in which it was published was judged eligible. After, another reviewer (LH) filled an spreadsheeted with a list of included and excluded journals. The search strategy is presented in the **Appendix**.

### Data extraction process

The included journals were uploaded to DistillerSR© by a library technician. We piloted 10% of the journals to tailor a standardized data extraction form (*repository link omitted for peer-reviewing purposes*). The variables were extracted independently by duplicates of reviewers (LH, DBR, FF, NA) and disagreements were resolved by consensus.

For each journal we identified its website *URL* and recorded variables of interest: (a) administrative details (e.g., publisher name); (b) bibliometric variables (e.g., 2017 JCR impact factor, DOAJPlus indexation); (c) openness (open-access level etc); (d) transparency (e.g., mention to a reporting guideline, EQUATOR Network, RCT registration requirement); (e) policy and guidance mentions of data sharing in the journal’s editorial processes and details; f) mechanisms and details of data-sharing statements whenever present, and mentions to IPD data sharing and their easy of accessibility. After our data extraction process, all journals were sent a copy of the completed data extraction for their respective journals and asked to verify or, if necessary, amend our records.

### Statistical Analysis

Statistical analyses were restricted to descriptive and a cross-tabulation of dichomitic variables. We calculated frequencies and estimated proportions with 95% confidence intervals for all categorical variables as well as median, interquartile range, and minimum and maximum values, excepted for pairwise cross-tabulations. These reulsts were displayed in a density plot. Analysis were done in Stata© v. 14.0.0 and R 3.4.1 (packages “*pacman*", “*tidyverse*” and “*reshape2*”) for MacOS.

### Data Sharing Statement and Plan

In addition to the protocol, our raw data (eligible and not eligible journals, Zotero library and dataset), statistical codes used in analysis, the glossary of variables, are available here (*repository link omitted for peer-reviewing purposes*). Any use of the data, or associated material, should be accompanied with their respective *digital object identifier* (DOI).

## Results

### Deviations from the protocol

We deviated from the protocol by removing the following journals from the project: Annals of Internal Medicine, JAMA, New England Journal of Medicine, BMJ, and Lancet. *A posteriori*, we perceived internal medicine journals as not reflecting Cardiology journals, which are specialty journals. Moreover, the NLM Catalog term “*Vascular Disease*” aforementioned in the methods was not planned at the moment of the first protocol draft.

### Main Results

Our search identified 450 journals. After de-duplication, 224 journals were included in our review. Twenty-three were excluded due to not having any publications in 2018, resulting in 201 potentially eligible journals. Another 50 journals were excluded as they did not publish any RCTs in 2018; one was excluded because of website inaccessibility; and two journals were not available in the eligible languages. Therefore, a total of 148 unique Cardiology journals were included. A flow diagram displays the flow of records/journals throughout the below (**Figure 1**).

**Figure 1.**
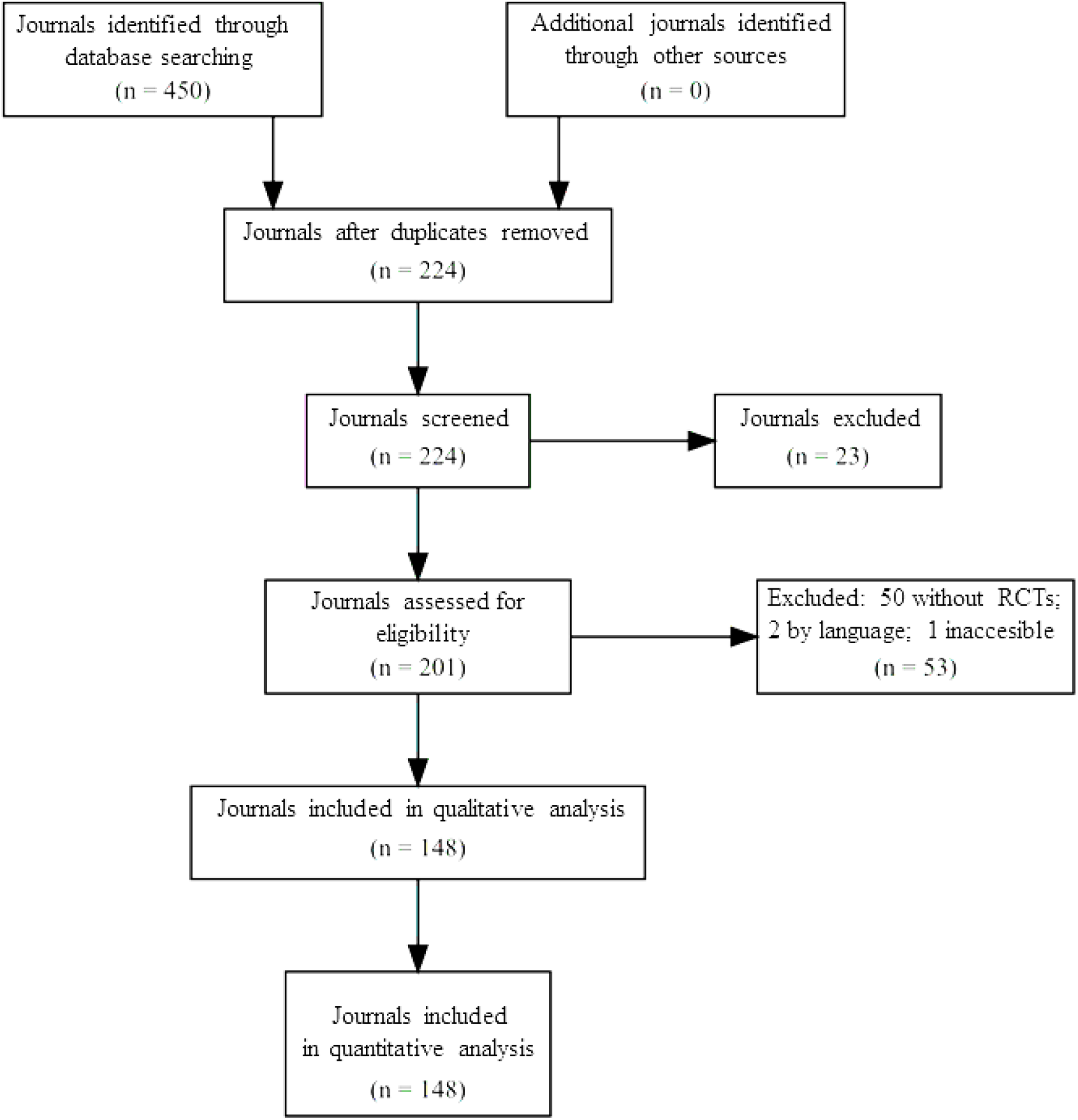
Flow Diagram of Included Journals.

Fifteen journals were indexed in DOAJPlus (10.1%, 95%CI 6.2% to 16.2%; *N* analyzed: 148), and 81 journals required RCTs registration in a public database (55.1%, 95%CI 46.9% to 63.0%; *N* analyzed: 147) (**Table 1)**.

**Table 1.** Journalology and reporting characteristics of evaluated journals.

| **Variables** | **Estimates** |
| --- | --- |
| **Publisher Type (N = 148)** |  |
| Academic | 3 (2.03%; 95%CI 6.46% to 61.7%) |
| Commercial | 145 (97.97%; 95% CI 93.82% to 99.35%) |
| **Open Access Type (N =148)** |  |
| Exclusively Subscription | 2 (1.35%; 95%CI 3.32% to 5.32%) |
| Hybrid | 134 (90.54%; 95%CI 84.57% to 94.35%) |
| Exclusively Open Access | 12 (8.11%; 95%CI 4.63% to 13.81%) |
| **DOAJPlus Indexing (N = 148)** |  |
| No | 133 (89.96%; 95%CI 83.78% to 93.83%) |
| Yes | 15 (10.14%; 95%CI 6.16% to 16.21%) |
| **JCR Impact Factor\* (N = 148)** |  |
|  | 2.73 (1.61 to 4.55; 0.36 to 23.42) |
| **RCT Registration Requirement (N = 147)** |  |
| No | 1 (0.68%; 95%CI 0.09% to 4.77%) |
| No explicit mention | 62 (42.18%; 95%CI 34.37% to 50.39%) |
| Encouragement | 3 (2.04%; 95%CI 0.65% to 6.21%) |
| Mandatory | 81 (55.10%; 95%CI 49.90% to 62.03%) |
| **Reporting Guidelines (N = 132)** |  |
| No | 64 (48.48%; 95%CI 39.97% to 57.08%) |
| Others | 5 (3.79%; 95%CI 1.56% to 8.87%) |
| Link to external source | 5 (3.79%; 95%CI 1.56% to 8.87%) |
| No CONSORT neither EQUATOR Network | 31 (23.48%; 95%CI 16.95% to 31.57%) |
| CONSORT with EQUATOR Network | 27 (20.45%; 95%CI 14.34% to 28.29%) |

**Notes:** 95%CI – 95% frequentist confidence interval; JCR Impact Factor\* - median (p25% to p75%; min – max); N – N analyzed.

For our primary outcome, 74 journals mention data sharing (whether being policy or guidance) (50.0%, 95%CI 41.9% to 58.1%; *N* analyzed = 148). When analyzing by policy or guidance, the results were similar. Six journals cited the need for a data-sharing statement in accordance with the ICMJE requirements^11^ (8.5%, 95%CI 3.8% to 17.9%; *N* analyzed: 71). Three journals encouraged data sharing but did not indicate a need to cite the ICMJE standards^11^ (4.2%, 95%CI, 0.8% to 12.0%; *N* analyzed: 70). In terms of the the ease of accessing the editorial policy on the journal website, our assessment considered 71 journals that had them easily accessible (97.2%, 95%CI 89.3% to 99.3%; *N* analyzed: 73). Two out of 68 journals had policies explicitly related to IPD data sharing (2.9%, 95%CI 0.7% to 11.4%), as displayed in **Table 2**. In the data-sharing guidance analysis, 56 journals disclosed whether a specific study design would be targeted. Eight provided data-sharing guidance specifically for RCTs (11.3%, 95%CI 5.6% to 21.3%). Sixty-five journals provided a mechanism for data sharing (95.6%, 95%CI 86.9% to 98.6%; *N* analyzed = 68). Importantly, out of 68 journals, only two provided important features guidance, particularly specific ones for IPD data sharing and to disclose who could request (or access) the data (*N* = 2 for both variables). Conversely, 67 disclosed the need to indicate the repository for components storage (98.5%, 95%CI 89.8% to 99.8%; *N* analyzed = 68). (**Table 2**).

**Table 2.** Study’s primary and secondary outcomes.

| **Variables** | **Estimates** |
| --- | --- |
| **Primary Outcome (N = 148)** |  |
| Composite Outcome | 74 (50%; 95%CI 41.90% to 58.10%) |
| Policies | 74 (50%; 95%CI 41.90% to 58.10%) |
| Guidance | 68 (47.50%; 95%CI 39.40% to 55.80%) |
| **Secondary Outcomes** |  |
| **Policies** |  |
| **Data-Sharing Statement Requirement (N = 73)** |  |
| Encouragement | 67 (91.78%; 95%CI 82.60 % to 96.33%) |
| Only cites the 2017 ICMJE Statement | 3 (4.11%; 95%CI 1.25% to 12.27%) |
| Mandatory | 3 (4.11%; 95%CI 1.25% to 12.27%) |
| **Policy Target (N = 73)** |  |
| None | 11 (15.07%; 95%CI 08.42% to 25.49%) |
| Experimental designs in general | 59 (80.82%; 95%CI 69.88% to 88.44%) |
| For RCTs and other designs | 1 (1.37%; 95%CI 1.83% to 9.48%) |
| Only for RCTs | 2 (2.74%; 95%CI 6.63% to 10.61%) |
| **Accessibility Level (N = 73)** |  |
| Hard to find | 2 (2.74%; 95%CI 0.66% to 10.61%) |
| Easy to find | 71 (97.26%; 95%CI 89.38% to 99.33%) |
| **Guidance** |  |
| **Guidance Target (N = 68)** |  |
| None | 6 (8.45%; 95%CI 3.77% to 17.86%) |
| Yes (unclear design) | 56 (78.87%; 95%CI 67.55% to 87.00%) |
| For RCTs and other designs | 8 (11.25%; 95%CI 5.63% to 21.25%) |
| Only for RCTs | 1 (1.41%; 95%CI 1.88% to 9.74%) |
| **How to share the data** |  |
| Unclear (N = 68) | 1 (1.45%; 95%CI 0.19% to 10.02%) |
| Materials (N = 63) | 59 (93.65%; 95%CI 83.89% to 97.66%) |
| Repository (N = 68) | 65 (95.59%; 95%CI 86.85% to 98.61%) |
| IPD data-sharing (N = 68) | 2 (2.94%; 95%CI 0.71% to 11.37%) |
| Who (N = 68) | 2 (2.94%; 95%CI 0.71% to 11.37%) |
| Availability Conditions (N = 67) | 61 (91.04%; 95%CI 81.12% to 96.00%) |

**Notes:** 95%CI – 95% frequentist confidence interval; “Guidance” is conditional to “Policy” variable. *N* – *N* analyzed.

Regarding instructions and guidance on how to report the clinical trials, 30 journals recommended or required using CONSORT with mention of the EQUATOR Network (20.5%, 95%CI 14.3% to 28.0%; *N* analyzed: 146); in turn, six journals did not mentioned anything about, and 38 journals made some recommendation without citing the CONSORT or EQUATOR Network. We also illustrated pairwise proportions between 10 variables related to a data-sharing plan in a density plot). We found that the highest association was between guidance on disclosure regarding data availability and the need to disclose the consiered repository. For IPD data sharing, low associations were found with all variables. (**Figure 2)**

**Figure 2.**
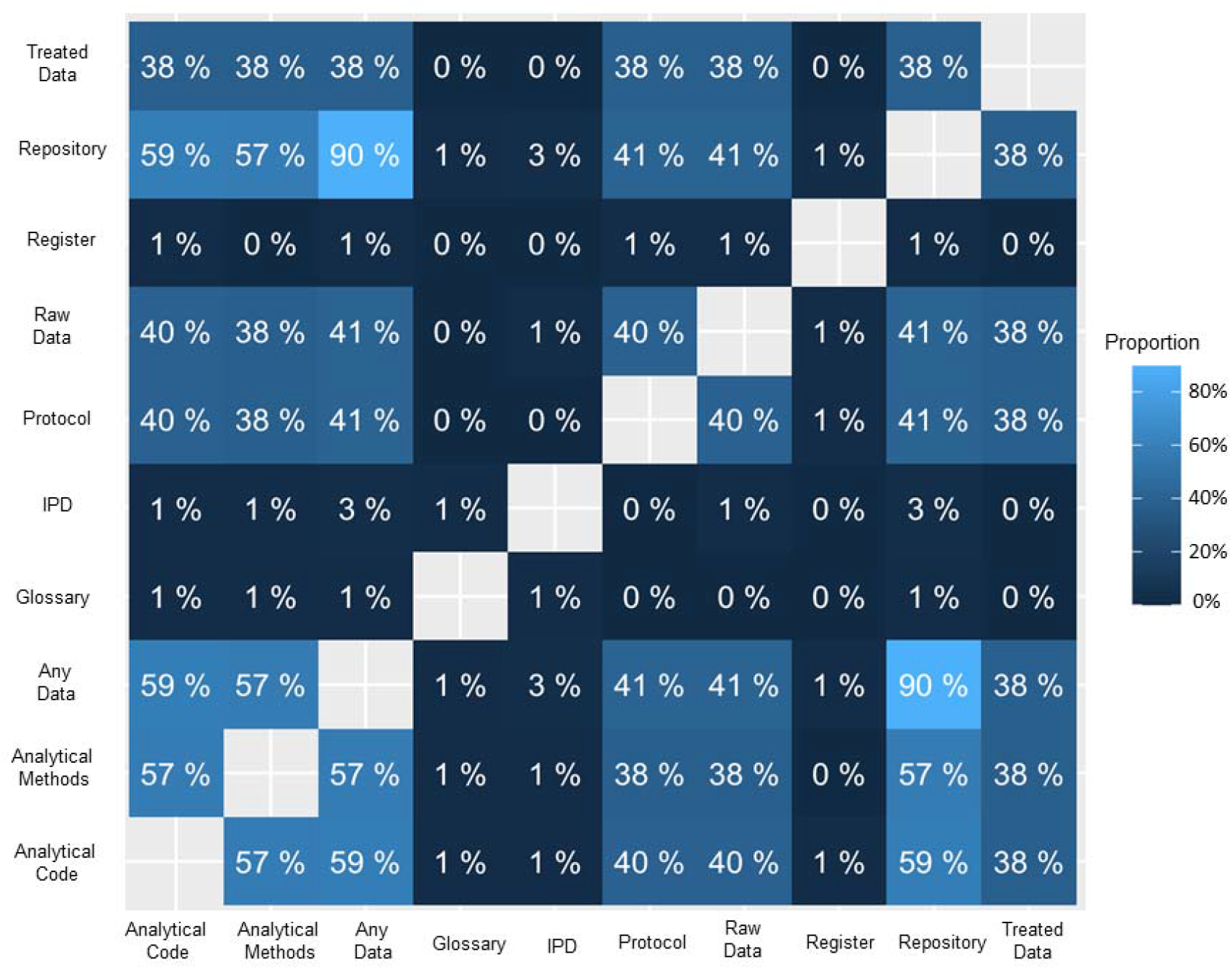
Pairwise cross-tabulation density plot between variables containing guidance herein.

## Discussion

Overall, we found the journals in our study had a mild adherence to our primary outcome, and these findings were consistent for policy and guidance analyzed separately, particularly with low adherence to IPD data sharing. Guidance for reporting clinical trials had a similar pattern, in which nearly half of the journals recommended or indicated the use of CONSORT, regardless of the presence of the EQUATOR Network for additional consultation. Moreover, we found a modest proportion for prospective registration as a condition for publication for RCTs in the evaluated journals. Concerning where and details for how to share the data, instructions were rarely reported, notably regarding the need to indicate the type of data considered for sharing. An exception is for the mention of repository details.

Primarily, data-sharing policy or guidance in their editorial contents were modest – nearly 50%, either being an independent mandatory policy or a reference about the ICMJE 2017 Statement. This does not seem to be unique to the Cardiology field. Resnik and colleagues^14^ found similar estimates for biomedical sciences; and the same for Computational Sciences within a very similar sample size of ours – ∼38% (*N* = 171);^15^ also closer to Biology and Microbiology (∼ 31%).^6^ Interestingly, Alsheikh-Ali and colleagues found that biomedical journals with higher impact factors are more prone to have data-sharing policies in their editorial contents^16^.

Guidance on the data-sharing process was rarely present. We need to acknowledge that we are plenty of external materials related to guidance on data-sharing – manuscripts, tutorials, recommendations of societies, independent third-parties initiatives that journals and editors can make use.^21^ The presence of the policy *plus guidance*, although not presumed to be a duty for the journal, could help to mitigate avoidable waste in research.

Impressingly, only two journals explicitly mentioned that IPD data sharing is highly encouraged. This is a cornerstone best practice in clinical research for a myriad of purposes and perhaps the core central message of our findings.^19^ The lack of data-sharing guidance, whenever the policy is present and the incipient adherence to them for IPD data sharing in particular are of high priority. On the other hand, the editorial contents for what materials should be shared and the necessity to deposit in a repository is more commonly stated.^20^

The transparency of a trial, mirrored by a trustworthy, open-access journal; transparent reporting; and prospective registration of the trial is crucial to the usability of policies and guidance for data sharing, likely accompaniying the existence of the practices.^22,23^ Our results found that only a few journals were indexed in DOAJPlus although claiming themselves as open access, and nearly half of them required prospective registration of randomized clinical trials, as above mentioned. Moreover, half of them did not mention anything about reporting guideline usage and, importantly, only 27 disclosed that CONSORT was recommended to report RCTs and that other reporting guidelines or extensions could be found in the EQUATOR Network website.

We should aknowledge our limitations. First, our list of variables can be considered somewhat subjective to the interpretation of the extractor given the inexistency of a lexicon for contents. Second, we limited our sample to journals indexed on the NLM Catalog and that have published at least one RCT in 2018. In addition, the term “IPD", could be hidden in the single term “data” in policies/guidance sections of journals, leading us to miss potentially eligible statements. Lastly, our list of variables was based on core items which illustrate policies and guidance for data-sharing based on the overall literature. An expanded investigation may provide questions we could not be able to answer.

On the other hand, we poise that our results should be considered by editors, funders and other stakeholders, potentially acting to drive research reproducibility and usability.^24,25^ Also, evidence-synthesis groups may benefit by IPD data for cumulative meta-analysis and IPD meta-analysis – otherwise, they would be unable to conduct it^26^; or we could go through basic arguments from philosophy of science, which state clearly that “*the knowledge is of no one*” and that data sharing should be an categorical imperative of researchers, once patients put themselves at risk whenever they are being enrolled in a trial^27,28^. There is too much to be done in all areas of research, but the Cardiology field, paradoxically, is filled by important, well-funded and well-designed trials that often change care decisions but lacks a lot on transparency and data-sharing policies. The private sponsorship, particularly industy funding, should incorporate practices herein; or, in another circumstances, regulatory agencies.

## Conclusion

In conclusion, Cardiology journals rarely mention data-sharing policies, transparency and reproducibility standards, and our empirical data supports a claim for the need for urgent improvement of such policies to mitigate this situation.

## Data Availability

As stated above, this research project works under a data and materials sharing regimen. Moreover, the results will be presented to third parties, send in an aggregated data manner to all journal editors previously screened and forwarded to interested stakeholders for potential improvements (such as the ICMJE, COPE, WAME and others) as well as to community overall.

## Ethical Approval & IRB

ethical approval for this study was exempted given no humans or animals involvement at any part.

## Acknowledgments

We acknowledge Mr. Raymond Daniel (the Ottawa Hospital Research Institute) for all the contributions with the library management and all editorial boards of included journals to provide feedback in the liability of our data-extraction.

## Contributions

LH, DU and DM conceived the study; LH, DBR, DU and DM refined study’s protocol; BS took care of all librarian activities; LH, FF, DBR and NA screened and confirmed eligibility of journals; LH, FF and DBR extracted the data; LH and FF tabulated results and made contact with journal editors for double checking; LH did all statistical analysis; LH wrote the first draft of the manuscript; all authors made substantial considerations to the following revisions and approved the final version of it.

## Appendix 1 Search Strategy

Final Strategy 2018

Dec 27

MEDLINE

Database: Ovid MEDLINE(R) and Epub Ahead of Print, In-Process & Other Non-Indexed Citations and Daily <1946 to December 26, 2018>

Search Strategy:

--------------------------------------------------------------------------------

1. randomized controlled trial.pt. (473462)
2. controlled clinical trial.pt. (92820)
3. randomized.ab. (430114)
4. placebo.ab. (194218)
5. clinical trials as topic.sh. (185597)
6. randomly.ab. (302507)
7. trial.ti. (191861)
8. or/1-7 (1189111)
9. exp animals/ not humans.sh. (4530051)
10. 8 not 9 (1093782)
11. limit 10 to yr=”2018” (63666)
12. acta cardiologica.jn. (4415)
13. acta myologica.jn. (311)
14. acute cardiac care.jn. (356)
15. advances in cardiology.jn. (1047)
16. american heart journal.jn. (24134)
17. “american journal of cardiology”.jn. (36653)
18. “american journal of cardiovascular drugs”.jn. (808)
19. “american journal of hypertension”.jn. (7185)
20. “american journal of physiology”.jn. (54733)
21. “anatolian journal of cardiology”.jn. (1125)
22. angiologiia i sosudistaia khirurgiia angiology & vascular surgery.jn. (1396)
23. angiology.jn. (7725)
24. annales de cardiologie et d angeiologie.jn. (3917)
25. “annals of cardiac anaesthesia”.jn. (1275)
26. “annals of noninvasive electrocardiology”.jn. (1290)
27. “annals of thoracic & cardiovascular surgery”.jn. (2061)
28. “annals of vascular surgery”.jn. (6304)
29. “archives of cardiovascular diseases”.jn. (1119)
30. archivos de cardiologia de mexico.jn. (1334)
31. arquivos brasileiros de cardiologia.jn. (8305)
32. arteriosclerosis thrombosis & vascular biology.jn. (8916)
33. asian cardiovascular & thoracic annals.jn. (2760)
34. atherosclerosis.jn. (13504)
35. atherosclerosis supplements.jn. (383)
36. basic research in cardiology.jn. (3492)
37. blood coagulation & fibrinolysis.jn. (3945)
38. blood pressure.jn. (1500)
39. blood pressure monitoring.jn. (1393)
40. bmc cardiovascular disorders.jn. (1839)
41. “brazilian journal of cardiovascular surgery”.jn. (292)
42. “canadian journal of cardiology”.jn. (6079)
43. cardiac electrophysiology clinics.jn. (461)
44. cardiology.jn. (4517)
45. cardiology clinics.jn. (1886)
46. “46 cardiology in review”.jn. (1006)
47. cardiology in the young.jn. (3693)
48. cardiology journal.jn. (1466)
49. cardiorenal medicine.jn. (267)
50. cardiovascular & interventional radiology.jn. (5736)
51. cardiovascular diabetology.jn. (1557)
52. cardiovascular drugs & therapy.jn. (2629)
53. cardiovascular engineering & technology.jn. (255)
54. cardiovascular & hematological agents in medicinal chemistry.jn. (355)
55. cardiovascular & hematological disorders drug targets.jn. (365)
56. “cardiovascular intervention and therapeutics”.jn. (555)
57. “cardiovascular journal of africa”.jn. (1217)
58. cardiovascular pathology.jn. (1549)
59. cardiovascular research.jn. (10628)
60. cardiovascular revascularization medicine.jn. (1399)
61. cardiovascular therapeutics.jn. (620)
62. cardiovascular toxicology.jn. (694)
63. cardiovascular ultrasound.jn. (701)
64. catheterization & cardiovascular interventions.jn. (8541)
65. cerebrovascular diseases.jn. (3071)
66. cerebrovascular diseases extra.jn. (142)
67. circulation.jn. (43168)
68. “circulation arrhythmia and electrophysiology”.jn. (1838)
69. circulation cardiovascular imaging.jn. (1512)
70. circulation cardiovascular interventions.jn. (1462)
71. circulation cardiovascular quality & outcomes.jn. (1295)
72. “circulation genomic and precision medicine”.jn. (151)
73. circulation heart failure.jn. (1416)
74. Circulation journal.jn. (6511)
75. circulation research.jn. (17193)
76. clinical & applied thrombosis hemostasis.jn. (1947)
77. clinical & experimental hypertension.jn. (192)
78. clinical cardiology.jn. (6339)
79. clinical hemorheology & microcirculation.jn. (2290)
80. clinical research in cardiology.jn. (1874)
81. clinical research in cardiology supplements.jn. (45)
82. clinica e investigacion en arteriosclerosis.jn. (268)
83. congenital heart disease.jn. (1295)
84. coronary artery disease.jn. (2766)
85. “critical pathways in cardiology a journal of evidence based medicine”.jn. (552)
86. current atherosclerosis reports.jn. (1475)
87. current cardiology reports.jn. (1795)
88. current cardiology reviews.jn. (469)
89. current heart failure reports.jn. (597)
90. current hypertension reports.jn. (1737)
91. current hypertension reviews.jn. (195)
92. current neurovascular research.jn. (607)
93. current opinion in cardiology.jn. (2441)
94. current opinion in nephrology & hypertension.jn. (2484)
95. Current problems in cardiology.jn. (614)
96. current vascular pharmacology.jn. (1131)
97. diabetes & vascular disease research.jn. (747)
98. echocardiography.jn. (4912)
99. esc heart failure.jn. (365)
100. eurointervention.jn. (3530)
101. europace.jn. (5210)
102. european heart journal.jn. (17292)
103. european heart journal acute cardiovascular care.jn. (606)
104. european heart journal cardiovascular imaging.jn. (2010)
105. european heart journal cardiovascular pharmacotherapy.jn. (211)
106. “european heart journal quality of care & clinical outcomes”.jn. (199)
107. “european journal of cardio thoracic surgery”.jn. (12260)
108. “european journal of cardiovascular nursing”.jn. (1001)
109. “european journal of heart failure”.jn. (3426)
110. “european journal of preventive cardiology”.jn. (1718)
111. “european journal of vascular & endovascular surgery”.jn. (6235)
112. “expert review of cardiovascular therapy”.jn. (2059)
113. future cardiology.jn. (1063)
114. general thoracic & cardiovascular surgery.jn. (1771)
115. giornale italiano di cardiologia.jn. (6827)
116. global heart.jn. (417)
117. harvard heart letter.jn. (1851)
118. heart.jn. (10152)
119. heart advisor.jn. (1129)
120. heart & vessels.jn. (2561)
121. heart failure clinics.jn. (772)
122. Heart failure reviews.jn. (973)
123. heart lung & circulation.jn. (2733)
124. heart & lung.jn. (4256)
125. heart rhythm.jn. (5542)
126. heart surgery forum.jn. (1751)
127. “hjc hellenic journal of cardiology”.jn. (1373)
128. herz.jn. (3289)
129. herzschrittmachertherapie und elektrophysiologie.jn. (854)
130. high blood pressure & cardiovascular prevention.jn. (362)
131. hipertension y riesgo vascular.jn. (137)
132. hypertension.jn. (14155)
133. hypertension in pregnancy.jn. (765)
134. hypertension research clinical & experimental.jn. (3495)
135. indian heart journal.jn. (5139)
136. innovations.jn. (40)
137. interactive cardiovascular & thoracic surgery.jn. (6457)
138. international angiology.jn. (2153)
139. international heart journal.jn. (1626)
140. “international journal of cardiology”.jn. (25551)
141. “international journal of stroke”.jn. (1949)
142. interventional cardiology clinics.jn. (368)
143. jacc cardiovascular imaging.jn. (2748)
144. jacc cardiovascular interventions.jn. (3812)
145. jacc clinical electrophysiology.jn. (752)
146. jacc heart failure.jn. (966)
147. jama cardiology.jn. (906)
148. journal de medecine vasculaire.jn. (95)
149. “journal of atherosclerosis & thrombosis”.jn. (1874)
150. “journal of cardiac failure”.jn. (2611)
151. “journal of cardiac surgery”.jn. (4187)
152. “journal of cardiology”.jn. (3761)
153. “journal of cardiopulmonary rehabilitation & prevention”.jn. (704)
154. “journal of cardiothoracic & vascular anesthesia”.jn. (6770)
155. “journal of cardiothoracic surgery”.jn. (1694)
156. “journal of cardiovascular computed tomography”.jn. (980)
157. “journal of cardiovascular electrophysiology”.jn. (6387)
158. “journal of cardiovascular magnetic resonance”.jn. (1380)
159. “journal of cardiovascular medicine”.jn. (2554)
160. “journal of cardiovascular nursing”.jn. (1847)
161. “journal of cardiovascular pharmacology”.jn. (11559)
162. “journal of cardiovascular pharmacology & therapeutics”.jn. (1093)
163. “journal of cardiovascular surgery”.jn. (7033)
164. “journal of cardiovascular translational research”.jn. (826)
165. “journal of cerebral blood flow & metabolism”.jn. (6142)
166. “journal of clinical hypertension”.jn. (3218)
167. “journal of echocardiography”.jn. (403)
168. “journal of electrocardiology”.jn. (4950)
169. “journal of endovascular therapy”.jn. (2533)
170. “journal of heart & lung transplantation”.jn. (6412)
171. “journal of heart valve disease”.jn. (3427)
172. “journal of human hypertension”.jn. (4508)
173. “journal of hypertension”.jn. (9769)
174. “journal of interventional cardiac electrophysiology”.jn. (2071)
175. “journal of interventional cardiology”.jn. (1771)
176. “journal of invasive cardiology”.jn. (4815)
177. “journal of molecular & cellular cardiology”.jn. (8129)
178. “journal of nuclear cardiology”.jn. (3968)
179. “journal of stroke & cerebrovascular diseases”.jn. (4546)
180. “journal of the american college of cardiology”.jn. (25113)
181. “journal of the american heart association”.jn. (3640)
182. “journal of the american society of hypertension”.jn. (1088)
183. “journal of thoracic & cardiovascular surgery”.jn. (26498)
184. “journal of thrombosis & haemostasis”.jn. (6520)
185. “journal of vascular access”.jn. (1492)
186. “journal of vascular & interventional radiology”.jn. (6893)
187. “journal of vascular nursing”.jn. (694)
188. “journal of vascular research”.jn. (1321)
189. “journal of vascular surgery”.jn. (15047)
190. j vasc surg venous lymphat disord.ja. (794)
191. “journal of veterinary cardiology”.jn. (583)
192. kardiologia polska.jn. (6764)
193. kardiologiia.jn. (13528)
194. methodist debakey cardiovascular journal.jn. (579)
195. microcirculation.jn. (1315)
196. microvascular research.jn. (3525)
197. minerva cardioangiologica.jn. (5861)
198. “multimedia manual of cardiothoracic surgery”.jn. (419)
199. nature reviews cardiology.jn. (1941)
200. nutrition metabolism & cardiovascular diseases.jn. (2277)
201. pacing & clinical electrophysiology.jn. (11111)
202. pediatric cardiology.jn. (5323)
203. perfusion.jn. (2019)
204. phlebology.jn. (1101)
205. pregnancy hypertension.jn. (1018)
206. progress in cardiovascular diseases.jn. (2187)
207. reviews in cardiovascular medicine.jn. (787)
208. revista espanola de cardiologia.jn. (8986)
209. revista portuguesa de cardiologia.jn. (3966)
210. revista portuguesa de cirurgia cardio toracica e vascular.jn. (745)
211. scandinavian cardiovascular journal.jn. (1371)
212. scandinavian cardiovascular journal supplement.jn. (33)
213. seminars in thoracic & cardiovascular surgery.jn. (1884)
214. seminars in thoracic & cardiovascular surgery pediatric cardiac surgery annual.jn. (384)
215. seminars in thrombosis & hemostasis.jn. (3115)
216. seminars in vascular surgery.jn. (932)
217. shock.jn. (4807)
218. stroke.jn. (19491)
219. techniques in vascular & interventional radiology.jn. (614)
220. texas heart institute journal.jn. (3766)
221. therapeutic advances in cardiovascular disease.jn. (381)
222. thoracic & cardiovascular surgeon reports.jn. (121)
223. thrombosis & haemostasis.jn. (13516)
224. thrombosis research.jn. (13042)
225. topics in stroke rehabilitation.jn. (1143)
226. translational stroke research.jn. (671)
227. trends in cardiovascular medicine.jn. (1767)
228. turk kardiyoloji dernegi arsivi.jn. (1877)
229. vasa.jn. (2946)
230. vascular.jn. (1210)
231. vascular & endovascular surgery.jn. (1708)
232. vascular health & risk management.jn. (1121)
233. vascular medicine.jn. (1343)
234. vascular pharmacology.jn. (1317)
235. world journal for pediatric & congenital heart surgery.jn. (1049)
236. zhonghua xin xue guan bing za zhi.ja. (4685)
237. or/12-236 [CARDIOLOGY JOURNALS – NLM CATALOG] (917693)
238. 11 and 12 [acta cardiologica] (3)
239. 11 and 13 [acta myologica] (0)
240. 11 and 14 [acute cardiac care] (0)
241. 11 and 15 [advances in cardiology] (0)
242. 11 and 16 [american heart journal] (113)
243. 11 and 17 [american journal of cardiology] (88)
244. 11 and 18 [american journal of cardiovascular drugs] (31)
245. 11 and 19 [american journal of hypertension] (17)
246. 11 and 20 [american journal of physiology] (0)
247. 11 and 21 [anatolian journal of cardiology] (8)
248. 11 and 22 [angiologiia i sosudistaia khirurgiia angiology & vascular surgery] (5)
249. 11 and 23 [angiology] (11)
250. 11 and 24 [annales de cardiologie et d angeiologie] (5)
251. 11 and 25 [annals of cardiac anaesthesia] (16)
252. 11 and 26 [annals of noninvasive electrocardiology] (7)
253. 11 and 27 [annals of thoracic & cardiovascular surgery] (3)
254. 11 and 28 [annals of vascular surgery] (22)
255. 11 and 29 [archives of cardiovascular diseases] (5)
256. 11 and 30 [archivos de cardiologia de mexico] (1)
257. 11 and 31 [arquivos brasileiros de cardiologia] (12)
258. 11 and 32 [arteriosclerosis thrombosis & vascular biology] (13)
259. 11 and 33 [asian cardiovascular & thoracic annals] (4)
260. 11 and 34 [atherosclerosis] (38)
261. 11 and 35 [atherosclerosis supplements] (0)
262. 11 and 36 [basic research in cardiology] (1)
263. 11 and 37 [blood coagulation & fibrinolysis] (4)
264. 11 and 38 [blood pressure] (7)
265. 11 and 39 [blood pressure monitoring] (7)
266. 11 and 40 [bmc cardiovascular disorders] (35)
267. 11 and 41 [”brazilian journal of cardiovascular surgery”] (12)
268. 11 and 42 [canadian journal of cardiology] (23)
269. 11 and 43 [cardiac electrophysiology clinics] (3)
270. 11 and 44 [cardiology] (8)
271. 11 and 45 [cardiology clinics] (2)
272. 11 and 46 [cardiology in review] (9)
273. 11 and 47 [cardiology in the young] (6)
274. 11 and 48 [cardiology journal] (22)
275. 11 and 49 [cardiorenal medicine] (2)
276. 11 and 50 [cardiovascular & interventional radiology] (17)
277. 11 and 51 [cardiovascular diabetology] (21)
278. 11 and 52 [cardiovascular drugs & therapy] (22)
279. 11 and 53 [cardiovascular engineering & technology] (0)
280. 11 and 54 [cardiovascular & hematological agents in medicinal chemistry] (1)
281. 11 and 55 [cardiovascular & hematological disorders drug targets] (5)
282. 11 and 56 [cardiovascular intervention and therapeutics] (4)
283. 11 and 57 [cardiovascular journal of africa] (1)
284. 11 and 58 [cardiovascular pathology] (0)
285. 11 and 59 [cardiovascular research] (9)
286. 11 and 60 [cardiovascular revascularization medicine] (47)
287. 11 and 61 [cardiovascular therapeutics] (16)
288. 11 and 62 [cardiovascular toxicology] (4)
289. 11 and 63 [cardiovascular ultrasound] (1)
290. 11 and 64 [catheterization & cardiovascular interventions] (88)
291. 11 and 65 [cerebrovascular diseases] (5)
292. 11 and 66 [cerebrovascular diseases extra] (0)
293. 11 and 67 [circulation] (127)
294. 11 and 68 [circulation arrhythmia and electrophysiology] (15)
295. 11 and 69 [circulation cardiovascular imaging] (3)
296. 11 and 70 [circulation cardiovascular interventions] (30)
297. 11 and 71 [circulation cardiovascular quality & outcomes] (24)
298. 11 and 72 [circulation genomic and precision medicine] (4)
299. 11 and 73 [circulation heart failure] (19)
300. 11 and 74 [Circulation journal] (33)
301. 11 and 75 [circulation research] (18)
302. 11 and 76 [clinical & applied thrombosis hemostasis] (15)
303. 11 and 77 [clinical & experimental hypertension] (0)
304. 11 and 78 [clinical cardiology] (46)
305. 11 and 79 [clinical hemorheology & microcirculation] (13)
306. 11 and 80 [clinical research in cardiology] (37)
307. 11 and 81 [clinical research in cardiology supplements] (0)
308. 11 and 82 [clinica e investigacion en arteriosclerosis] (4)
309. 11 and 83 [congenital heart disease] (5)
310. 11 and 84 [coronary artery disease] (19)
311. 11 and 85 [critical pathways in cardiology] (3)
312. 11 and 86 [current atherosclerosis reports] (6)
313. 11 and 87 [current cardiology reports] (19)
314. 11 and 88 [current cardiology reviews] (5)
315. 11 and 89 [current heart failure reports] (5)
316. 11 and 90 [current hypertension reports] (9)
317. 11 and 91 [current hypertension reviews] (5)
318. 11 and 92 [current neurovascular research] (1)
319. 11 and 93 [current opinion in cardiology] (18)
320. 11 and 94 [current opinion in nephrology & hypertension] (6)
321. 11 and 95 [current problems in cardiology] (2)
322. 11 and 96 [current vascular pharmacology] (14)
323. 11 and 97 [diabetes & vascular disease research] (7)
324. 11 and 98 [echocardiography] (5)
325. 11 and 99 [ESC heart failure] (38)
326. 11 and 100 [eurointervention] (47)
327. 11 and 101 [europace] (50)
328. 11 and 102 [european heart journal] (80)
329. 11 and 103 [european heart journal acute cardiovascular care] (13)
330. 11 and 104 [european heart journal cardiovascular imaging] (12)
331. 11 and 105 [european heart journal cardiovascular pharmacotherapy] (10)
332. 11 and 106 [european heart journal quality of care & clinical outcomes] (6)
333. 11 and 107 [european journal of cardio thoracic surgery] (26)
334. 11 and 108 [european journal of cardiovascular nursing] (19)
335. 11 and 109 [european journal of heart failure] (54)
336. 11 and 110 [european journal of preventive cardiology] (26)
337. 11 and 111 [european journal of vascular & endovascular surgery] (21)
338. 11 and 112 [expert review of cardiovascular therapy] (13)
339. 11 and 113 [future cardiology] (4)
340. 11 and 114 [general thoracic & cardiovascular surgery] (7)
341. 11 and 115 [giornale italiano di cardiologia] (9)
342. 11 and 116 [global heart] (2)
343. 11 and 117 [harvard heart letter] (0)
344. 11 and 118 [heart] (21)
345. 11 and 119 [heart advisor] (0)
346. 11 and 120 [heart & vessels] (20)
347. 11 and 121 [heart failure clinics] (3)
348. 11 and 122 [heart failure reviews] (18)
349. 11 and 123 [heart lung & circulation] (15)
350. 11 and 124 [heart & lung] (8)
351. 11 and 125 [heart rhythm] (24)
352. 11 and 126 [heart surgery forum] (3)
353. 11 and 127 [HJC hellenic journal of cardiology] (9)
354. 11 and 128 [herz] (31)
355. 11 and 129 [herzschrittmachertherapie und elektrophysiologie] (5)
356. 11 and 130 [high blood pressure & cardiovascular prevention] (7)
357. 11 and 131 [hipertension y riesgo vascular] (1)
358. 11 and 132 [hypertension] (27)
359. 11 and 133 [hypertension in pregnancy] (6)
360. 11 and 134 [hypertension research clinical & experimental] (9)
361. 11 and 135 [indian heart journal] (12)
362. 11 and 136 [innovations] (0)
363. 11 and 137 [interactive cardiovascular & thoracic surgery] (37)
364. 11 and 138 [international angiology] (11)
365. 11 and 139 [international heart journal] (13)
366. 11 and 140 [international journal of cardiology] (126)
367. 11 and 141 [international journal of stroke] (48)
368. 11 and 142 [interventional cardiology clinics] (5)
369. 11 and 143 [jacc cardiovascular imaging] (30)
370. 11 and 144 [jacc cardiovascular interventions] (65)
371. 11 and 145 [jacc clinical electrophysiology] (15)
372. 11 and 146 [jacc heart failure] (27)
373. 11 and 147 [jama cardiology] (38)
374. 11 and 148 [journal de medecine vasculaire] (3)
375. 11 and 149 [journal of atherosclerosis & thrombosis] (17)
376. 11 and 150 [journal of cardiac failure] (17)
377. 11 and 151 [journal of cardiac surgery] (6)
378. 11 and 152 [journal of cardiology] (24)
379. 11 and 153 [journal of cardiopulmonary rehabilitation & prevention] (24)
380. 11 and 154 [journal of cardiothoracic & vascular anesthesia] (66)
381. 11 and 155 [journal of cardiothoracic surgery] (8)
382. 11 and 156 [journal of cardiovascular computed tomography] (5)
383. 11 and 157 [journal of cardiovascular electrophysiology] (19)
384. 11 and 158 [journal of cardiovascular magnetic resonance] (4)
385. 11 and 159 [journal of cardiovascular medicine] (8)
386. 11 and 160 [journal of cardiovascular nursing] (13)
387. 11 and 161 [journal of cardiovascular pharmacology] (14)
388. 11 and 162 [journal of cardiovascular pharmacology & therapeutics] (17)
389. 11 and 163 [journal of cardiovascular surgery] (24)
390. 11 and 164 [journal of cardiovascular translational research] (7)
391. 11 and 165 [journal of cerebral blood flow & metabolism] (9)
392. 11 and 166 [journal of clinical hypertension] (23)
393. 11 and 167 [journal of echocardiography] (0)
394. 11 and 168 [journal of electrocardiology] (5)
395. 11 and 169 [journal of endovascular therapy] (11)
396. 11 and 170 [journal of heart & lung transplantation] (15)
397. 11 and 171 [journal of heart valve disease] (0)
398. 11 and 172 [journal of human hypertension] (14)
399. 11 and 173 [journal of hypertension] (49)
400. 11 and 174 [journal of interventional cardiac electrophysiology] (25)
401. 11 and 175 [journal of interventional cardiology] (21)
402. 11 and 176 [journal of invasive cardiology] (18)
403. 11 and 177 [journal of molecular & cellular cardiology] (1)
404. 11 and 178 [journal of nuclear cardiology] (8)
405. 11 and 179 [journal of stroke & cerebrovascular diseases] (62)
406. 11 and 180 [journal of the american college of cardiology] (83)
407. 11 and 181 [journal of the american heart association] (85)
408. 11 and 182 [journal of the american society of hypertension] (13)
409. 11 and 183 [journal of thoracic & cardiovascular surgery] (58)
410. 11 and 184 [journal of thrombosis & haemostasis] (28)
411. 11 and 185 [journal of vascular access] (21)
412. 11 and 186 [journal of vascular & interventional radiology] (20)
413. 11 and 187 [journal of vascular nursing] (1)
414. 11 and 188 [journal of vascular research] (0)
415. 11 and 189 [journal of vascular surgery] (61)
416. 11 and 190 [j vasc surg venous lymphat disord] (15)
417. 11 and 191 [journal of veterinary cardiology] (0)
418. 11 and 192 [kardiologia polska] (8)
419. 11 and 193 [kardiologiia] (8)
420. 11 and 194 [methodist debakey cardiovascular journal] (2)
421. 11 and 195 [microcirculation] (2)
422. 11 and 196 [microvascular research] (6)
423. 11 and 197 [minerva cardioangiologica] (14)
424. 11 and 198 [multimedia manual of cardiothoracic surgery] (0)
425. 11 and 199 [nature reviews cardiology] (5)
426. 11 and 200 [nutrition metabolism & cardiovascular diseases] (22)
427. 11 and 201 [pacing & clinical electrophysiology] (25)
428. 11 and 202 [pediatric cardiology] (9)
429. 11 and 203 [perfusion] (17)
430. 11 and 204 [phlebology] (15)
431. 11 and 205 [pregnancy hypertension] (9)
432. 11 and 206 [progress in cardiovascular diseases] (13)
433. 11 and 207 [reviews in cardiovascular medicine] (1)
434. 11 and 208 [revista espanola de cardiologia] (12)
435. 11 and 209 [revista portuguesa de cardiologia] (7)
436. 11 and 210 [revista portuguesa de cirurgia cardio toracica e vascular] (0)
437. 11 and 211 [scandinavian cardiovascular journal] (4)
438. 11 and 212 [scandinavian cardiovascular journal supplement] (0)
439. 11 and 213 [seminars in thoracic & cardiovascular surgery] (8)
440. 11 and 214 [seminars in thoracic & cardiovascular surgery pediatric cardiac surgery annual] (0)
441. 11 and 215 [seminars in thrombosis & hemostasis] (4)
442. 11 and 216 [seminars in vascular surgery] (0)
443. 11 and 217 [shock] (47)
444. 11 and 218 [stroke] (90)
445. 11 and 219 [techniques in vascular & interventional radiology] (0)
446. 11 and 220 [texas heart institute journal] (1)
447. 11 and 221 [therapeutic advances in cardiovascular disease] (4)
448. 11 and 222 [thoracic & cardiovascular surgeon reports] (0)
449. 11 and 223 [thrombosis & haemostasis] (21)
450. 11 and 224 [thrombosis research] (31)
451. 11 and 225 [topics in stroke rehabilitation] (19)
452. 11 and 226 [translational stroke research] (9)
453. 11 and 227 [trends in cardiovascular medicine] (5)
454. 11 and 228 [turk kardiyoloji dernegi arsivi] (2)
455. 11 and 229 [vasa] (11)
456. 11 and 230 [vascular] (7)
457. 11 and 231 [vascular & endovascular surgery] (7)
458. 11 and 232 [vascular health & risk management] (6)
459. 11 and 233 [vascular medicine] (7)
460. 11 and 234 [vascular pharmacology] (3)
461. 11 and 235 [world journal for pediatric & congenital heart surgery] (3)
462. 11 and 236 [zhonghua xin xue guan bing za zhi] (10)

* * * * * * * * * * * * * * * * * * * * * * * * * * *

## References

1. Ross JS, Madigan D, Hill KP, Egilman DS, Wang Y, Krumholz HM. Pooled analysis of rofecoxib placebo-controlled clinical trial data: lessons for postmarket pharmaceutical safety surveillance. Arch Intern Med. Nov 23 2009;169(21):1976–85. doi:10.1001/archinternmed.2009.394

2. Riley RD, Lambert PC, Abo-Zaid G. Meta-analysis of individual participant data: rationale, conduct, and reporting. BMJ. 2010;340:c221. doi:10.1136/bmj.c221

3. Moylan EC, Kowalczuk MK. Why articles are retracted: a retrospective cross-sectional study of retraction notices at BioMed Central. BMJ Open. Nov 23 2016;6(11):e012047. doi:10.1136/bmjopen-2016-012047

4. Gay HC, Baldridge AS, Huffman MD. Feasibility, Process, and Outcomes of Cardiovascular Clinical Trial Data Sharing: A Reproduction Analysis of the SMART-AF Trial. JAMA Cardiology. Dec 1 2017;2(12):1375–1379. doi:10.1001/jamacardio.2017.3808.

5. Vaduganathan M, Nagarur A, Qamar A, et al. Availability and Use of Shared Data From Cardiometabolic Clinical Trials. Circulation. 2018/02/27 2018;137(9):938–947. doi:10.1161/CIRCULATIONAHA.117.031883.

6. Vasilevsky NA, Minnier J, Haendel MA, Champieux RE. Reproducible and reusable research: are journal data sharing policies meeting the mark? PeerJ. 2017;5:e3208. doi:10.7717/peerj.3208.

7. Taichman DB, Sahni P, Pinborg A, et al. Data Sharing Statements for Clinical Trials: A Requirement of the International Committee of Medical Journal Editors. Ann Intern Med. 2017:63–65.

8. Nosek BA, Alter G, Banks GC, et al. Promoting an open research culture. Science. 2015;348(6242):1422. doi:10.1126/science.aab2374.

9. Iqbal SA, Wallach JD, Khoury MJ, Schully SD, Ioannidis JP. Reproducible Research Practices and Transparency across the Biomedical Literature. PLoS Biol. Jan 2016;14(1):e1002333. doi:10.1371/journal.pbio.1002333.

10. Wallach JD, Boyack KW, Ioannidis JPA. Reproducible research practices, transparency, and open access data in the biomedical literature, 2015-2017. PLoS Biol. Nov 2018;16(11):e2006930. doi:10.1371/journal.pbio.2006930.

11. Collaboration OS. PSYCHOLOGY. Estimating the reproducibility of psychological science. Science. Aug 2015;349(6251):aac4716. doi:10.1126/science.aac4716.

12. von Elm E, Altman DG, Egger M, et al. The Strengthening the Reporting of Observational Studies in Epidemiology (STROBE) statement: guidelines for reporting observational studies. J Clin Epidemiol. 2008;61(4):344–349. doi:10.1016/j.jclinepi.2007.11.008.

13. Dickersin K, Scherer R, Lefebvre C. Identifying relevant studies for systematic reviews. BMJ. Nov 12 1994;309(6964):1286–91. doi:10.1136/bmj.309.6964.1286.

14. Resnik DB, Morales M, Landrum R, et al. Effect of impact factor and discipline on journal data sharing policies. Account Res. 2019/04/03 2019;26(3):139–156. doi:10.1080/08989621.2019.1591277.

15. Stodden V, Guo P, Ma Z. Toward Reproducible Computational Research: An Empirical Analysis of Data and Code Policy Adoption by Journals. PLoS One. 2013;8(6):e67111. doi:10.1371/journal.pone.0067111.

16. Alsheikh-Ali AA, Qureshi W, Al-Mallah MH, Ioannidis JPA. Public Availability of Published Research Data in High-Impact Journals. PLoS One. 2011;6(9):e24357. doi:10.1371/journal.pone.0024357.

17. Kidwell MC, Lazarević LB, Baranski E, et al. Badges to Acknowledge Open Practices: A Simple, Low-Cost, Effective Method for Increasing Transparency. PLoS Biol. 2016;14(5):e1002456. doi:10.1371/journal.pbio.1002456.

18. Boutron I, Dechartres A, Baron G, Li J, Ravaud P. Sharing of Data From Industry-Funded Registered Clinical Trials. JAMA. 2016;315(24):2729–2730. doi:10.1001/jama.2016.6310.

19. Ohmann C, Banzi R, Canham S, et al. Sharing and reuse of individual participant data from clinical trials: principles and recommendations. BMJ Open. 2017;7(12):e018647. doi:10.1136/bmjopen-2017-018647.

20. Banzi R, Canham S, Kuchinke W, Krleza-Jeric K, Demotes-Mainard J, Ohmann C. Evaluation of repositories for sharing individual-participant data from clinical studies. Trials. 2019/03/15 2019;20(1):169. doi:10.1186/s13063-019-3253-3;

21. Wilkinson MD, Dumontier M, Aalbersberg IJ, et al. The FAIR Guiding Principles for scientific data management and stewardship. Scient Data. 2016/03/15 2016;3(1):160018. doi:10.1038/sdata.2016.18.

22. Turner L, Shamseer L, Altman DG, et al. Consolidated standards of reporting trials (CONSORT) and the completeness of reporting of randomised controlled trials (RCTs) published in medical journals. Cochrane Database of Systematic Reviews. 2012;(11)doi:10.1002/14651858.MR000030.pub2.

23. Munafò MR, Nosek BA, Bishop DVM, et al. A manifesto for reproducible science. Nature Human Behav. 2017/01/10 2017;1(1):0021. doi:10.1038/s41562-016-0021.

24. Ioannidis JPA. How to Make More Published Research True. PLOS Med. 2014;11(10):e1001747. doi:10.1371/journal.pmed.1001747.

25. Ebrahim S, Sohani ZN, Montoya L, et al. Reanalyses of Randomized Clinical Trial Data. JAMA. 2014;312(10):1024–1032. doi:10.1001/jama.2014.9646.

26. Rogozinska E, Marlin N, Thangaratinam S, Khan KS, Zamora J. Meta-analysis using individual participant data from randomised trials: opportunities and limitations created by access to raw data. Evid Based Med. Oct 2017;22(5):157–162. doi:10.1136/ebmed-2017-110775.

27. Hardwicke TE, Mathur MB, MacDonald K, et al. Data availability, reusability, and analytic reproducibility: evaluating the impact of a mandatory open data policy at the journal Cognition. R Soc Open Sci. Aug 2018;5(8):180448. doi:10.1098/rsos.180448.

28. Rowhani-Farid A, Barnett AG. Has open data arrived at the British Medical Journal (BMJ)? An observational study. BMJ Open. 2016;6(10):e011784. doi:10.1136/bmjopen-2016-011784.

